# Economic precarity, social isolation, and suicidal ideation during the COVID-19 pandemic

**DOI:** 10.1101/2020.10.05.20205955

**Authors:** Julia Raifman, Catherine K. Ettman, Lorraine Dean, Colleen Barry, Sandro Galea

## Abstract

**Importance:** The US population faces stressors associated with suicide brought on by the COVID-19 pandemic. Understanding the relationship between stressors and suicidal ideation may inform policies and programs to prevent suicide.

**Objective:** To evaluate the relationship between stressors and suicidal ideation during the COVID-19 pandemic.

**Design:** We compared suicidal ideation in 2017-2018 to suicidal ideation in 2020. We estimated the association between stressors and suicidal ideation in bivariable and multivariable Poisson regression models with robust variance.

**Setting:** United States

**Participants:** Participants were from two, nationally representative surveys of US adults: The 2017-2017 National Health and Nutrition Examination Survey and the 2020 COVID-19 and Life Stressors Impact on Mental Health and Well-being study (conducted March 31 to April 13), analyzed April 28 to September 30, 2020.

**Exposures:** Economic precarity as measured through job loss or difficulty paying rent and social isolation based on reporting “feeling alone.”

**Main outcome measure:** Suicidal ideation based on reporting “Thoughts that you would be better off dead or of hurting yourself in some way” over the past two weeks.

**Results:** Suicidal ideation increased more than fourfold, from 3.4% in the 2017-2018 NHANES to 16.3% in the 2020 CLIMB survey, and from 5.8% to 26.4% among participants in low-income households. Suicidal ideation was more prevalent among people facing difficulty paying rent (31.5%), job loss (24.1%), and loneliness (25.1%), with each stressor associated with suicidal ideation in bivariable models. In the multivariable model, difficulty paying rent was associated with suicidal ideation (aPR: 1.5, 95% CI: 1.2 to 2.1), while losing a job was not (aPR: 0.9, 95% CI: 0.6 to 1.2). Feeling alone was associated with suicidal ideation (aPR: 1.9, 95% CI: 1.5 to 2.4).

**Conclusions and relevance:** Suicidal ideation increased more than fourfold during the COVID-19 pandemic. Difficulty paying rent and loneliness were most associated with suicidal ideation. Policies and programs to support people experiencing economic precarity and loneliness may contribute to suicide prevention.

---

During the coronavirus 2019 (COVID-19) pandemic the population of the United States (US) is facing several co-occurring stressors. The response to the pandemic has led to economic downturn, creating stressors of job loss and financial distress. Physical distancing to prevent the spread of COVID-19 occasions stressors including social isolation. Economic precarity^1^ and social isolation^2^ are associated with mental distress and suicide outside of the pandemic context. The prolonged intersection of these forces during the COVID-19 pandemic could have profound implications for suicide, already a leading cause of premature death in the US.^3^ Increases in suicide have occurred with prior pandemics,^4^ and emerging evidence of increased depression during the COVID-19 pandemic further raises concern about the risk for suicide during COVID-19.^5,6^ While suicide mortality data are typically not publicly available for several months after the end of each year, understanding the populations most at risk of suicidal ideation and the association between COVID-19 stressors and suicidal ideation can inform policies and programs to prevent suicide.

## Methods

### Sample

We used data from a nationally representative sample of US adults aged 18 or older collected through the AmeriSpeak standing panel. Panelists were invited to participate in the COVID-19 and Life Stressors Impact on Mental Health and Well-being (CLIMB) study from March 31, 2020 through April 13, 2020 and paid a cash equivalent of $3 for completing the survey. Of those invited to participate in the survey who had responded to a survey in the past 6 months, 64% completed it. As a pre-pandemic comparison, we used data from the 2017-2018 National Health and Nutrition Examination Survey (NHANES), a nationally representative sample of noninstitutionalized civilian US adults aged 18 years or older collected by the US government. The CLIMB and NHANES samples are comparable in that they are both nationally representative. We excluded participants who did not respond to questions about suicidal ideation in NHANES and participants who did not respond to any variables included in analyses in CLIMB data.

### Exposures

We evaluated three COVID-19 stressors reflecting economic precarity and social isolation, each measured as binary variables reported in response to a question, “Have any of the following affected your life as a result of the coronavirus or COVID-19 outbreak?” First, we measured job loss based on checking “losing a job.” Second, we measured difficulty paying rent based on checking “having difficulty paying rent.” Third, we measured social isolation as checking “feeling alone.”

### Outcome

We measured suicidal ideation based on Patient Health Questionnaire-9 (PHQ-9) item 9, which asks participants to rate the frequency with which they have had “Thoughts that you would be better off dead or of hurting yourself in some way” over the past two weeks and response options of “Not at all, several days, more than half the days, or nearly every day.” We created a binary variable for reporting these feelings with any frequency over the past two weeks. Prior research indicates responses to this question were correlated with future suicide attempts and deaths.^7–9^

### Analysis

First, we described the demographic characteristics of participants in the 2020 CLIMB data and in the 2017-2018 NHANES data. Second, we estimated the prevalence of suicidal ideation by demographic characteristics and calculated the share with suicidal ideation within subgroups in 2020 relative to 2017-2018. Third, we estimated unadjusted and adjusted prevalence ratios (PR and aPR) of the association between COVID-19 related stressors and suicidal ideation using a Poisson regression model with robust variance to approximate a log-binomial regression model, with α = 0.05.^10^ In the multivariable model, we adjusted for age group, education level, sex, race and ethnicity, household income, savings, marital status, COVID-19 illness, and COVID-19 bereavement.

## Results

A total of 1,415 (96.3%) of 1,470 CLIMB participants responded to all questions relevant to the analysis and 5,085 (86.8%) of 5,856 NHANES participants responded to suicidal ideation questions and were included in the samples. Demographic characteristics of both samples were nationally representative (**Table 1**). Overall, suicidal ideation increased more than fourfold, from 3.4% in the 2017-2018 NHANES to 16.3% in the 2020 CLIMB survey. The greatest absolute increases in suicidal ideation and 2020 prevalence of suicidal ideation were among participants earning less than $20,000 (5.8% to 26.4%), participants aged 18 to 29 (4.1% to 23.5%), and participants who were Hispanic (3.7% to 23.1%). In 2020, suicidal ideation was high among those who faced difficulty paying rent (31.5%, **Figure 1**) and job loss (24.1%), as well as loneliness (25.1%).

**Table 1:** Participant demographic characteristics, COVID-19 stressors, and suicidal ideation.

|  | Sample characteristics |  |  |  | Suicidal ideation |  |  |  | Absolute difference<br>2020 -<br>(2017-2018) | Ratio,<br>2020:<br>2017-2018 |
| --- | --- | --- | --- | --- | --- | --- | --- | --- | --- | --- |
|  | NHANES,<br>2017-2018 |  | CLIMB, 2020 |  | NHANES, 2017-2018, Suicidal<br>ideation |  | CLIMB, 2020 |  |  |  |
|  | n | % | n | % | n | % | n | % | Percentage<br>points |  |
| Total | 5,085 | 100 | 1,415 | 100 | 192 | 3.4 | 231 | 16.3 | 12.9 | 4.8 |
| Difficulty paying rent |  |  |  |  |  |  |  |  |  |  |
| No | N/A | N/A | 1,199 | 84.7 | N/A | N/A | 163 | 13.6 | N/A | N/A |
| Yes | N/A | N/A | 216 | 15.3 | N/A | N/A | 68 | 31.5 | N/A | N/A |
| Lost job |  |  |  |  |  |  |  |  |  |  |
| No | N/A | N/A | 1,253 | 88.5 | N/A | N/A | 192 | 15.3 | N/A | N/A |
| Yes | N/A | N/A | 162 | 11.5 | N/A | N/A | 39 | 24.1 | N/A | N/A |
| Feeling alone |  |  |  |  |  |  |  |  |  |  |
| No | N/A | N/A | 952 | 67.3 | N/A | N/A | 115 | 12.1 | N/A | N/A |
| Yes | N/A | N/A | 463 | 32.7 | N/A | N/A | 116 | 25.1 | N/A | N/A |
| Age group |  |  |  |  |  |  |  |  |  |  |
| 18-29 | 965 | 21.3 | 238 | 16.8 | 38 | 4.1 | 56 | 23.5 | 19.1 | 5.7 |
| 30-44 | 1,106 | 24.2 | 493 | 34.8 | 38 | 3.1 | 92 | 18.7 | 15.4 | 6.0 |
| 45-59 | 1,183 | 26.5 | 337 | 23.8 | 48 | 3.0 | 47 | 14 | 11 | 4.7 |
| 60+ | 1,831 | 28.1 | 347 | 24.5 | 68 | 3.5 | 36 | 10.4 | 6.8 | 2.9 |
| Sex |  |  |  |  |  |  |  |  |  |  |
| Male | 2,489 | 48.7 | 708 | 50 | 102 | 4.0 | 109 | 15.4 | 11.5 | 3.9 |
| Female | 2,596 | 51.3 | 707 | 50 | 90 | 2.9 | 122 | 17.3 | 13.9 | 5.8 |
| Race/ethnicity |  |  |  |  |  |  |  |  |  |  |
| Non-Hispanic White | 1,801 | 63 | 922 | 65.2 | 71 | 3.3 | 124 | 13.5 | 10.2 | 4.1 |
| Non-Hispanic Black | 1,178 | 11.1 | 137 | 9.7 | 33 | 3.1 | 22 | 16.1 | 12.1 | 4.9 |
| Non-Hispanic another race or<br>multiracial | 947 | 10.1 | 105 | 7.4 | 34 | 4.0 | 27 | 25.7 | 20.1 | 6.0 |
| Hispanic | 1,159 | 15.8 | 251 | 17.7 | 54 | 3.7 | 58 | 23.1 | 19.1 | 6.2 |
| Education level |  |  |  |  |  |  |  |  |  |  |
| High school graduate or less | 2,262 | 39.4 | 327 | 23.1 | 112 | 4.2 | 68 | 20.8 | 16.1 | 4.8 |
| Some college | 1,640 | 30.4 | 631 | 44.5 | 60 | 4.3 | 101 | 16 | 11.8 | 3.7 |
| College grad or more | 1,177 | 30.2 | 457 | 32.3 | 20 | 1.4 | 62 | 13.6 | 11.7 | 9.4 |
| Not reported | 6 | <0.1 | 0 | N/A | N/A | N/A | N/A | N/A | N/A | N/A |
| Marital status |  |  |  |  |  |  |  |  |  |  |
| Unmarried or separated | 2,424 | 45.1 | 716 | 50.6 | 127 | 4.6 | 152 | 21.2 | 16.4 | 4.6 |
| Married | 2,412 | 51.5 | 699 | 49.4 | 54 | 2.2 | 79 | 11.3 | 8.9 | 5.0 |
| Not reported | 249 | 3.4 | 0 | N/A | 11 | 5.2 | N/A | N/A | N/A | N/A |
| Household income |  |  |  |  | N/A | N/A |  |  |  |  |
| \$0-\$19,999 | 875 | 11.5 | 220 | 15.6 | 50 | 5.8 | 58 | 26.4 | 19.8 | 4.4 |
| \$20,000-\$39,999^ | 1,322 | 21.4 | 321 | 22.7 | 63 | 5.0 | 56 | 17.5 | 12.1 | 3.4 |
| \$40,000-\$74,999^ | 891 | 17.7 | 410 | 29 | 32 | 3.3 | 59 | 14.4 | 11.1 | 4.4 |
| ≥\$75,000 | 1,356 | 38.7 | 464 | 32.8 | 25 | 2.1 | 58 | 12.5 | 10.5 | 6.0 |
| Not reported | 641 | 10.7 | 0 | N/A | 22 | 2.7 | N/A | N/A | N/A | N/A |
| Savings |  |  |  |  |  |  |  |  |  |  |
| <\$5,000 | N/A | N/A | 720 | 50.9 | N/A | N/A | 155 | 21.5 | N/A | N/A |
| ≥\$5,000 | N/A | N/A | 695 | 49.1 | N/A | N/A | 76 | 10.9 | N/A | N/A |
| COVID-19 illness |  |  |  |  |  |  |  |  |  |  |
| No | N/A | N/A | 1,403 | 99.1 | N/A | N/A | 223 | 15.9 | N/A | N/A |
| Yes | N/A | N/A | 12 | 0.9 | N/A | N/A | 8 | 66.7 | N/A | N/A |
| Death of someone close due to<br>COVID-19 |  |  |  |  |  |  |  |  |  |  |
| No | N/A | N/A | 1,390 | 98.2 | N/A | N/A | 222 | 16 | N/A | N/A |
| Yes | N/A | N/A | 25 | 1.8 | N/A | N/A | 9 | 36 | N/A | N/A |

Notes: \*p<0.1, \*\*p<0.05, \*\*\*p<0.01. Percents are weighted. ^Income categories for NHANES participants were \$20,000 to \$44,999 and \$45,000 to \$74,999 based on different cut points for income questions. Data on suicidal ideation among those with COVID-19 illness and bereavement are based on small sample sizes.

**Figure 1:**
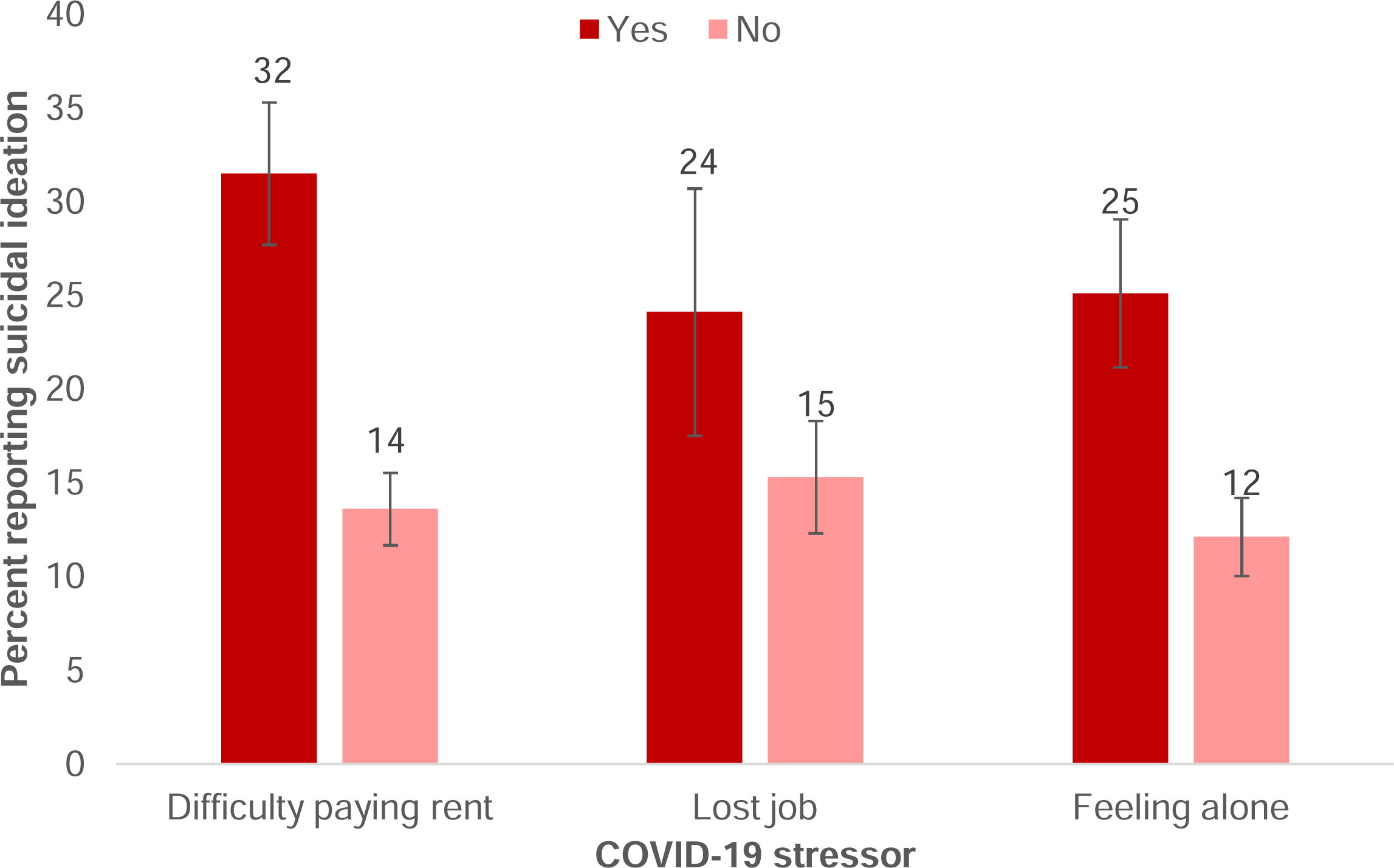
COVID-19 stressors and suicidal ideation.

Each of the stressors we evaluated were associated with suicidal ideation in the bivariable model (**Table 2**; Difficulty paying rent PR: 2.3, 95% CI: 1.8 to 3.1; feeling alone PR: 2.1, 95% CI: 1.6 to 2.6; job loss PR: 1.6, 95% CI: 1.1 to 2.2). In the multivariable model, difficulty paying rent was associated with suicidal ideation (aPR: 1.5, 95% CI: 1.2 to 2.1), while losing a job was not (aPR: 0.9, 95% CI: 0.6 to 1.2). Feeling alone was also associated with suicidal ideation (aPR: 1.9, 95% CI: 1.5 to 2.4). Although the sample size for persons with COVID-19 illness (n=12) or bereavement (n=25) is small, the results (66.7% and 36.0%, respectively) are suggestive that COVID-19 illness or bereavement may be associated with increased suicidal ideation.

**Table 2:**
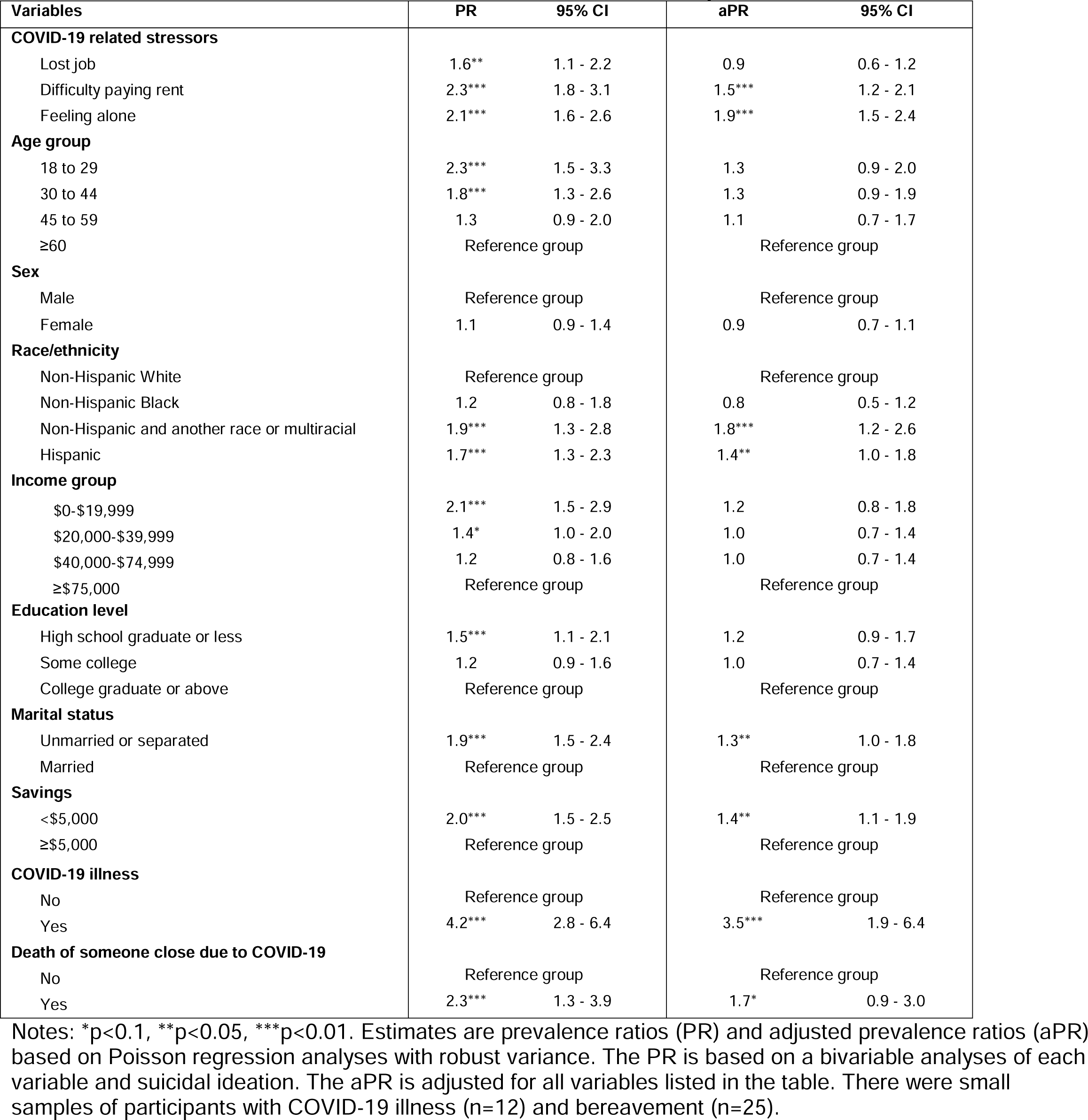
COVID-19 related stressors and suicidal ideation over the past two weeks (N=1415)

## Discussion

We found that there was a more than fourfold increase in suicidal ideation during the COVID-19 pandemic; 16.1% of people reported suicidal ideation, relative to 2017-2018, when 3.4% of people reported suicidal ideation. In keeping with prior studies on psychological distress and depression, we found that people living in low-income households and young people are particularly at risk of suicidal ideation during the COVID-19 pandemic.^5,6^

Reporting difficulty paying rent was associated with suicidal ideation. Prior research indicates that financial distress, such as that which has become widespread during the COVID-19 pandemic, is associated with suicide^1^ and that eviction in particular is associated with suicide.^11^ Policies such as the Center for Disease Control and Prevention’s federal eviction moratorium, state eviction moratoriums, and federal and state unemployment insurance policies^12^ and the federal stimulus payments may play help prevent suicide. While job loss was not associated with suicidal ideation during the CLIMB study period of late March and early April, it is important to study mental health among people who lost work due to the pandemic over the long term as high unemployment has been prolonged for several months, especially for people in low-income households and for people who are Black and Hispanic.^13^

People who reported feeling lonely were nearly twice as likely to report suicidal ideation, highlighting the need for programs and policies to provide social support, such as through social connections in environments with lower COVID-19 risk (e.g. outdoors) or via computer or phone.

This study was conducted in late March and early April 2020, when COVID-19 spread across the US was still in its early stages; as such we did not have a large enough sample of persons who had contracted COVID-19 or had loved ones who died to study the association between COVID-19 illness or bereavement with suicidal ideation. The results suggest a potential association between these exposures and suicidal ideation that warrants further study.

Finally, prior studies indicate that means restriction,^14^ particularly of firearms,^15^ is associated with reductions in suicide. Policies or programs to reduce household firearm ownership could play an important role in suicide prevention in the COVID-19 context of elevated stressors.

Limitations include that suicidal ideation was based on self-report and that the characteristics of participants in CLIMB and NHANES differed. Those who responded to surveys may have differed from those who did not, particularly if stressors affected survey participation. The CLIMB study was conducted early in the pandemic period, and the relationship between stressors and suicidal ideation may have changed as the pandemic and associated stressors continue to affect the US population.

## Conclusion

Suicidal ideation increased substantially during the COVID-19 pandemic. Those facing difficulty paying rent and loneliness may be at particular risk of suicide. Policies and programs to support people experiencing economic precarity and difficulty paying rent may contribute to suicide prevention, as may programs to support individuals facing prolonged social isolation.

## Data Availability

The NHANES data are publicly available. Those interested in the CLIMB data may email Ms. Ettman with inquiries.

https://www.cdc.gov/nchs/nhanes/index.htm

